# Application of Bayesian spatial modelling to uncover geographical disparities and improve antimicrobial resistant surveillance

**DOI:** 10.1101/2024.11.06.24316846

**Authors:** Teresa Maria Wozniak, Alys Young, David Conlan, Aminath Shausan, Amalie Dyda, Benn Sartorius, Marcela Cespedes

**Affiliations:** Australian e-Health Research Centre CSIRO, Herston, Queensland, Australia; Australian e-Health Research Centre CSIRO, Parkville, Victoria, Australia; School of Public Health, University of Queensland, Herston, Queensland, Australia; University of Queensland Centre for Clinical Research (UQCCR), The University of Queensland, Brisbane, Queensland, Australia

**Keywords:** disease surveillance, antimicrobial resistance, modelling, spatial epidemiology

## Abstract

**Introduction:** Disease surveillance is an essential element of an effective response to antimicrobial resistance (AMR). Associations between AMR cases and area-level drivers such as remoteness and socio-economic disadvantage have been observed, but spatial associations when modelling routinely collected surveillance data that are often imperfect or missing have not been previously possible.

**Aim:** We aimed to use spatial modelling to adjust for area-level variables and to enhance AMR surveillance for missing or sparse data, in an effort to provide clinicians and policy makers with more actionable epidemiological information.

**Methods:** We used monthly antimicrobial susceptibility data for methicillin-resistant *Staphylococcus aureus* (MRSA) from a surveillance system in Australia. MRSA was assessed for the effects of age, sex, socio-economic and access to healthcare services indices by fitting Bayesian spatial models.

**Results:** We analysed data for 77, 760 MRSA isolates between 2016 and 2022. We observed significant spatial heterogeneity in MRSA and found significant associations with age, sex and remoteness, but not socio-economic status. MRSA infections were highest in adult females aged 16-60 living in very remote regions and lowest in senior males aged 60+ years living in inner regional areas..

**Conclusion:** Current disease surveillance approaches for antimicrobial resistant infections have limited spatial comparability, are not timely, and at risk of sampling bias. Bayesian spatial models borrow information from neighbouring regions to adjust for unbalanced geographical information and can fill information gaps of current MRSA surveillance. Assessment of disease spatial variation is especially critical in settings which have diverse geography, dispersed populations or in regions with limited microbiological capacity.

## Introduction

One of the greatest global health challenges we face is the increase in antibiotic-resistant bacteria. In 2019, antimicrobial resistance (AMR) was directly responsible for 1.27 million deaths across the globe and increasing levels of disability and costs^1^. Australia has rising rates of AMR and comparatively high rates of antimicrobial use^2^. Remote regions of Australia, where the population density is low have some of the highest rates of AMR in the world, with 46% of *Staphylococcus aureus* isolates methicillin-resistant (MRSA)^3^. These regions have historically fallen outside of surveillance reach^4^, which may be in part due limited healthcare services and microbiology capacity dispersed across a large geographically diverse area and high staff turn over^4^^5^. Despite these surveillance challenges, healthcare professions are reliant on accessing reliable, local and timely data that represents the geographical diversity of the patient population that they serve^5^. Mapping disease patterns is one way to support these health practitioner information needs and to gain a better understanding of the interconnectedness of AMR across One Health ecosystem^6^.

Currently, mapping disease patterns from AMR surveillance systems is largely done using thematic maps^7,8^, as either a choropleth, heat map, plot densities, or cartogram. Such maps are often highly dependent on testing rates and may mislead the viewer into thinking an important pattern exists, when noise or sampling error are likely explanations^9,10^. In geographically diverse regions such as Europe, where population density and geographic characteristics are varied^11^, or in remote settings of Australia, with small sample size or outliers, these data are at risk of being disproportionately represented in thematic maps^12^.

Bayesian spatial statistics is a powerful approach to quantify disease patterns that takes into account information from surrounding areas (and in time), leveraging that nearby observations are often more likely to be similar ^13–15^. Taking advantage of this dependence across space (and time) allows one to smooth out the data, helping to counteract potential effects from outlier observations or uneven data structures/sample size, often seen in areas with low population or diverse geography. These models also more accurately quantify the significance of complex relationships between disease and key risk factors and can include a wide range of data sources, such as location, time, age groups, and gender ^16^.

Bayesian models are increasingly being used to enhance public health surveillance data, for identification of disease clusters ^17^, to correct reporting delays ^18^ and for epidemic tracking ^19^. However, these methods have been under-utilised for enhancing AMR surveillance for more timely detection of incident cases in regions with sparse data.

This study expands on earlier AMR mapping efforts in northern Australia ^3^ and contributes to the importance of using statistical approaches when transforming surveillance data into actionable epidemiological information.

## Materials and Methods

### Study setting

This study was set across three jurisdictions of northern Australia: Western Australia, Northern Territory and Queensland, which is divided into 22 Statistical Area level 3 (SA3) regions (Figure 1) ^12^.

**Figure 1.**
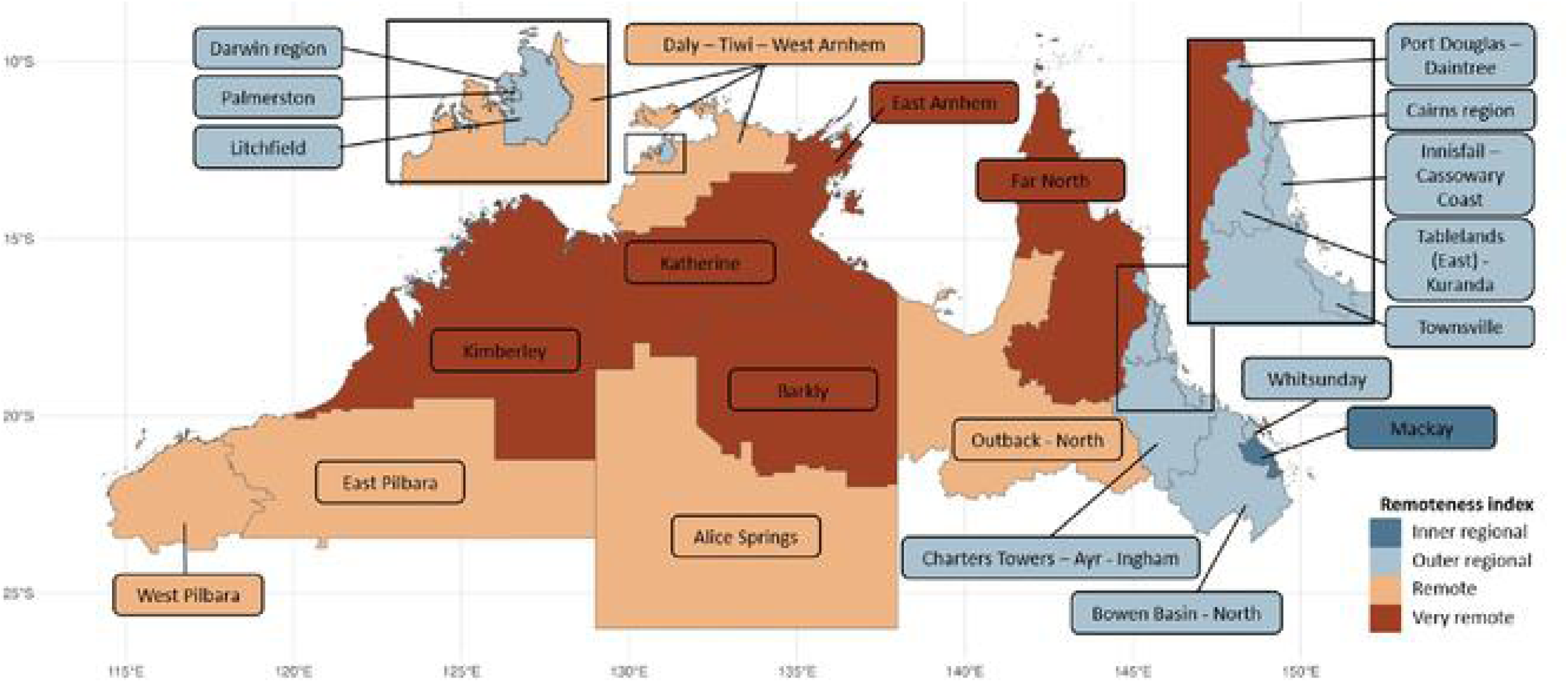
Study sites in northern Australia comprised of 22 regions colour coded by Remoteness index.

Northern Australia comprises of 50% ‘outer regional’, 23% remote and very remote and the remining 4% inner regional (**Figure 1**). The disadvantaged areas tended to be localised to regional and remote settings, while more advantaged areas tend to be in or near the regional cities **(Figure 2)**.

**Figure 2.**
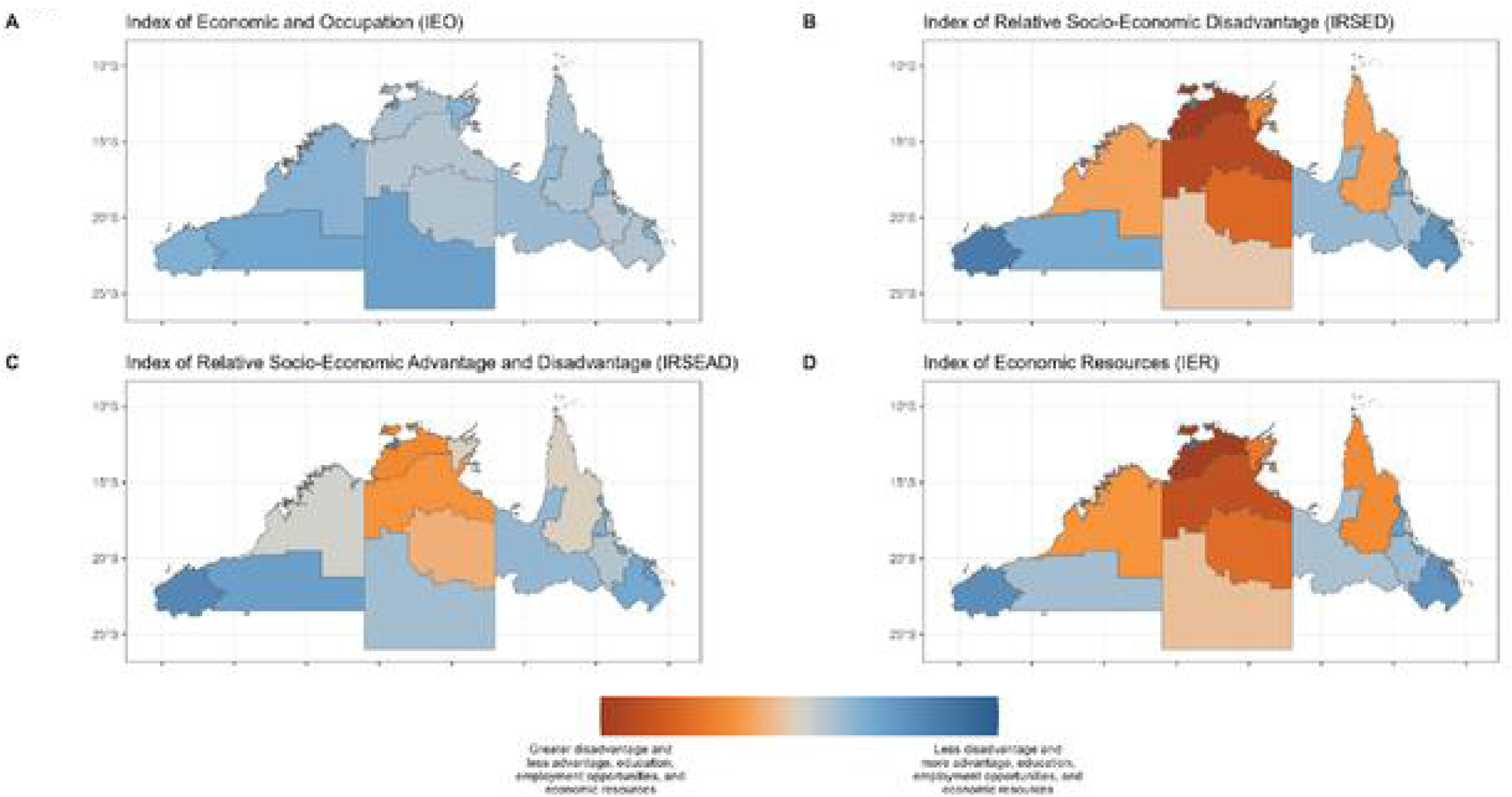
The socio-economic factors by region, northern Australia between 2016 and 2021. (A) Index of Economic and Occupation. (B) Index of Relative Socio-economic Disadvantage. (C) Index of Relative Socio-economic Advantage and Disadvantage. (D) Index of Economic Resources.

### Data source

We used surveillance data from the HOTspots surveillance and response program, which has been previously described^3^. Monthly methicillin-resistant *Staphylococcus aureus* (MRSA) isolates from hospitals and primary healthcare clinics was extracted for the years 2016 – 2022. Isolate information was recorded on 22 non-overlapping aerial regions for Northen Australia across three jurisdictions at the Statistical Area level 3 level, which is a regional breakdown of Australia with a population estimate of 30,000 to 130,000^12^. Age was aggregated to children (0-15 years), adults (16-60 years) and senior adults (61 years and over). Sex was assessed as male or female.

Remoteness index was classified into five categories (major cities, inner regional, outer regional, remote and very remote) ^12^.

Socio-Economic Indexes for Areas (SEIFA), includes four indices derived from the Australian Bureau of Statistics Census during the study period^20^. Namely, index of relative socio-economic advantage and disadvantage; index of relative socio-economic disadvantage; index of education and occupation; and index of economic resources.

### Statistical analyses

We analysed MRSA data at each time point for spatial autocorrelation via Moran’s I, which ranges from minus one to one. Moran’s I of minus one, denotes observations are perfectly spatially displaced (i.e. such as in a checkerboard); Moran’s I equal to zero, denotes observations resemble random patterns, and Moran’s I towards one suggests strong spatial autocorrelation and the potential presence of spatial clusters. Bayesian intrinsic conditional autoregressive (CAR) spatial models were fitted on MRSA prevalence (assuming a binomial distribution) independently at each time point which had significant spatial autocorrelation (Supplementary Figure 1). To avoid multicollinearity each SEIFA index was investigated separately and the index providing the best fit was included in the final multivariable model (Supplementary Figure 2).

MRSA prevalence was derived from the number of antibiotic susceptibility tests that were resistant (𝑟_𝑘_) divided by the number of total susceptibility tests (𝑡_𝑘_), for region 𝑘 at a specific time point. To investigate spatial autocorrelation and the effect of age and sex, as well as region specific Socio-Economic Indexes for Areas (SEIFA) and remoteness index on MRSA, the following model was fitted:

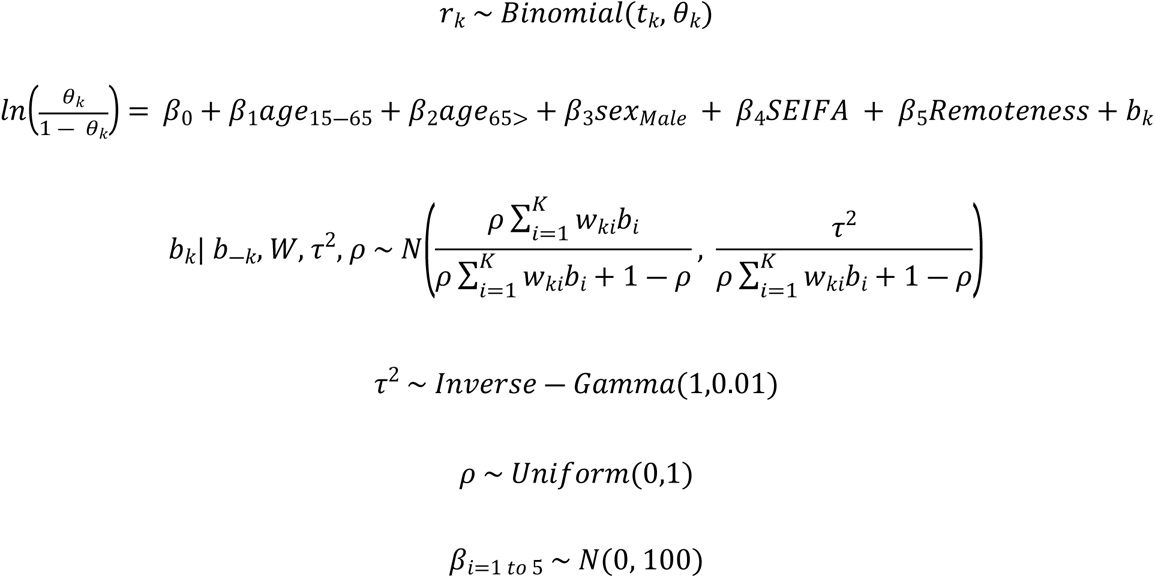

The logistic linear predictor comprises of the sum of the effects of age, sex, remoteness index and SEIFA indices in addition to the spatial autocorrelation, modelled via the random the spatial effects. These random effects follow the specification by Leroux et. al. (2000) ^21^ to apply a flexible approach to varying strengths of spatial autocorrelation.

The strength of spatial dependence, denoted by 𝜌, can range from strong and clustered spatial structure as 𝜌 approaches one, to independent and identically distributed observations, when 𝜌 equals zero. The spatial scale variance parameter, 𝜏^2^, describes the amount of variation between the spatial random effects. Incorporation of information to the neighbourhood structure is denoted by binary and symmetric matrix 𝑊. Geographical regions 𝑘 and 𝑙 which share a border with each other are denoted by 𝑤_𝑘𝑙_ = 1 and zero otherwise. Vague non-informative priors were chosen for parameters 𝜏^2^, 𝜌 and 𝛽. Estimation of joint posterior distribution was computed via Markov chain Monte Carlo (MCMC) methods^22^.

Ten models were explored at each time point (Supplementary Table 1), and convergences were visually assessed via density, trace and autocorrelation plots (Supplementary Table 2). The prevalence resistance with 95% credible intervals (Cr. I) for each region were estimated via posterior predictions.

For assessing the odds of MRSA infection by age, adult (aged 16 -60) and senior adult (aged 61+) categories were compared to the children (age 0-15). For assessing the odds of MRSA by remoteness, we compared all remoteness categories to inner regional.

All analyses were completed using R software version 4.2.1 (R Core Team 2022) within the RStudio editor (version 2022.07.02). The following R packages were used for data cleaning, processing, Moran’s I assessment, Bayesian model fitting, geographical maps and plots: “dplyr”, “reshape2”, “zoo”. “ggplot2”, “ggfortify”, “coda”, “spdep” and “sf”, “CARBayes”. All maps were generated using the ‘ggplot2’ and “sf”, packages ^23^.

### Ethical statement

This study was conducted and approved by the Human Research Ethics Committee of the Northern Territory Department of Health and Menzies School of Health Research (HREC-2018-3084). The study did not recruit active participants and used retrospective pathology records that were provided to the researchers in a deidentified format, a waiver of consent was approved.

## Results

### Prevalence of MRSA in northern Australia

The study utilised data on 77, 760 clinical isolates collected between 2016 and 2022 (84 months).

Figure 3 presents a descriptive analysis of MRSA, including healthcare onset and sex. According to the descriptive summary from the HOTspots surveillance program, MRSA prevalence has remained stable at 35% since 2016 albeit, there was a reduction in 2021 to 31% (**Figure 3A**). There was an observed difference in MRSA by onset, with higher prevalence of MRSA observed in community healthcare settings compared to hospitals since 2014 (**Figure 3B**). MRSA has been consistently higher amongst females compared to males (**Figure 3C**).

**Figure 3.**
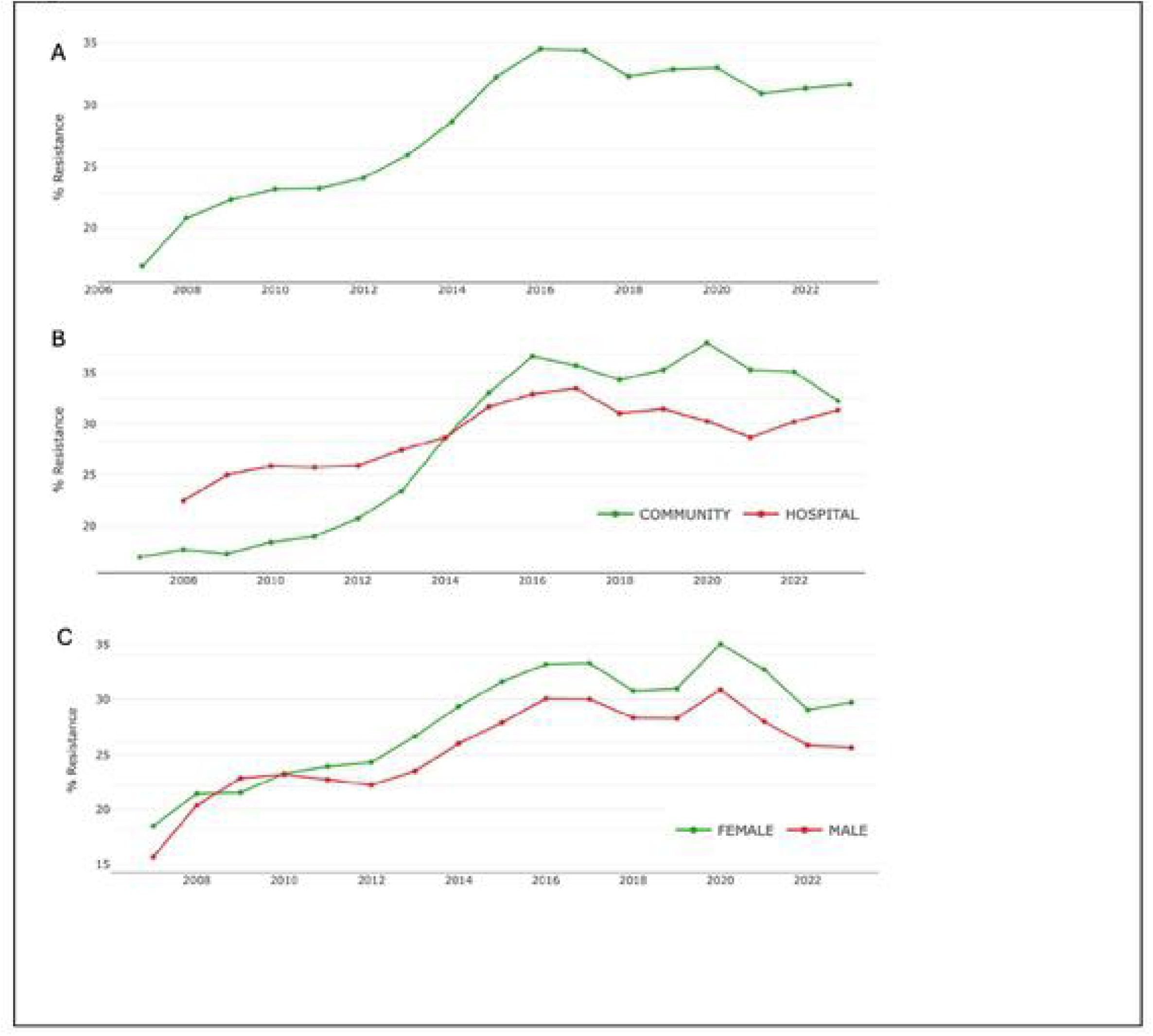
Epidemiology of MRSA, over time (A), by healthcare setting (B), and by sex (C), northern Australia 2008-2023 Source: Images generated from HOTspots digital surveillance platform https://amr-hotspots.net/ (accessed March 2024)

### Spatial autocorrelation and clusters

Figure 4A shows the results from the spatial autocorrelation analysis measured by Moran’s I index and the associated significance test. The analysis suggest high spatial heterogeneity with Moran’s I index ranging from −0.25 to 0.75, across regions between January 2016 until December 2022.

**Figure 4A.**
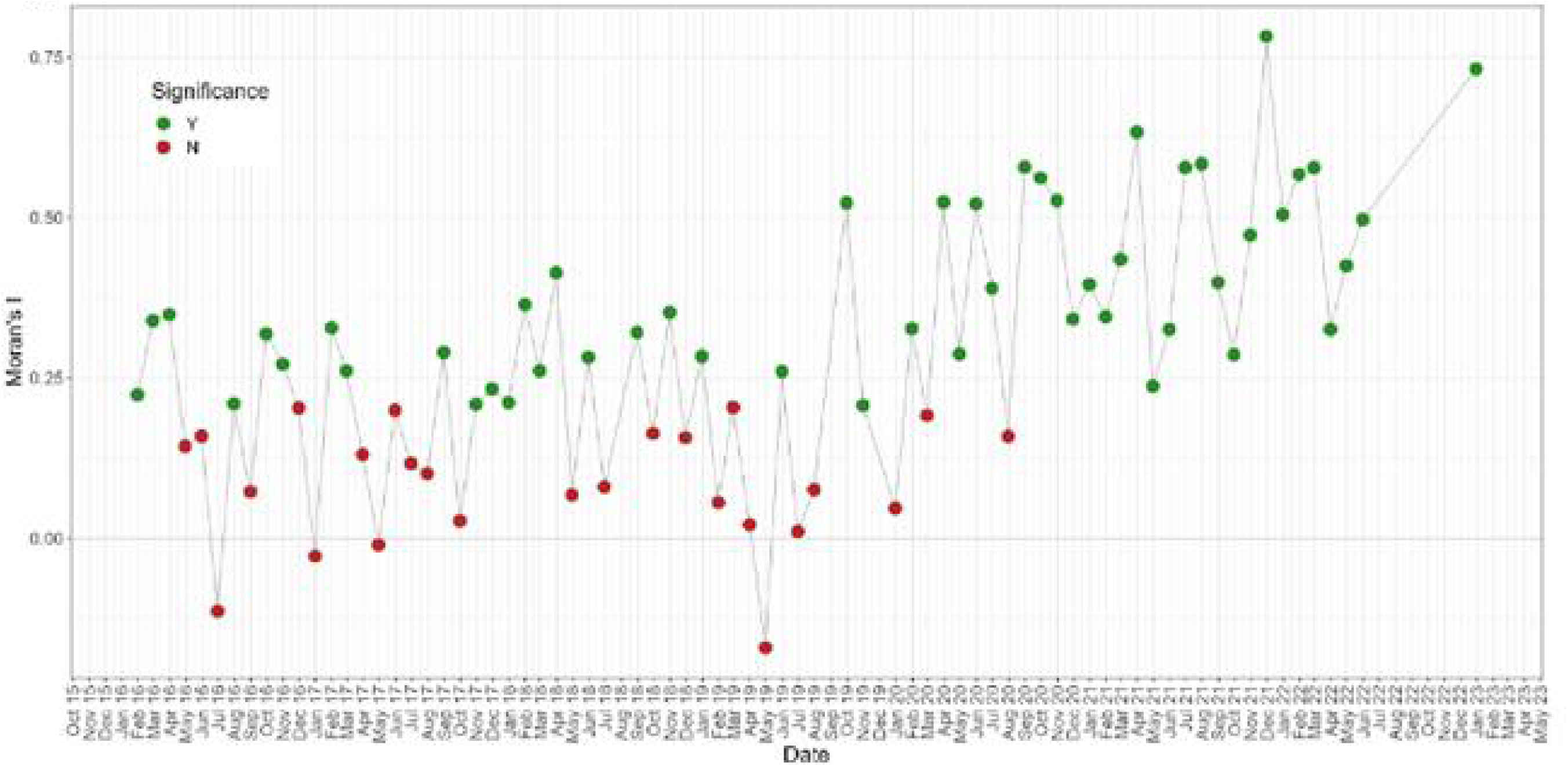
Spatial autocorrelation (Moran’s I statistic) of MRSA, northern Australia 2016-2022

Large fluctuations in spatial autocorrelation were observed for MRSA from January 2016 until September 2020. From October 2020, Moran’s I index was increasing reaching a maximum of 0.75 (December 2021, p < 0.001), indicating a stronger spatial autocorrelation and potential for cluster detection. Highest MRSA infections were observed in central regions of northern Australia, namely Pilbara (Western Australia), Kimberly (Western Australia), Barkly (Northern Territory), Alice Springs (Northern Territory) and Outback North (Queensland). Whereas clusters of low MRSA infections were observed in the top end of Northern Territory (Darwin region and the capital city of Northern Territory), and on the east coastline of northern Australia (Queensland) (Figure 4B).

**Figure 4B.**
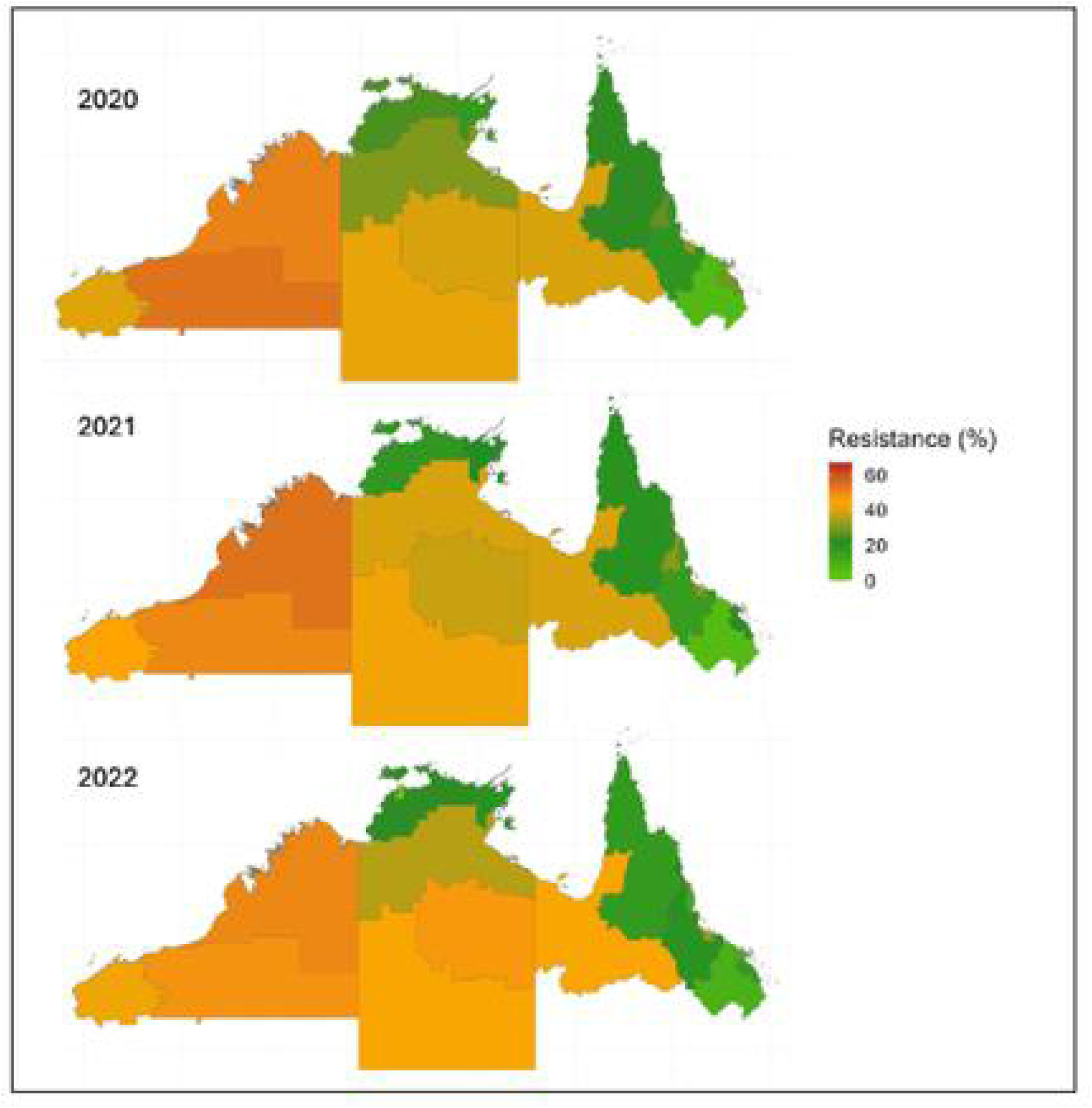
Regions of clustered high and low MRSA, northern Australia 2020-2022

### Spatial risk of MRSA by sex and age

Females exhibited a higher likelihood of MRSA infection compared to males, with an average odds ratio (OR) of 1.33 (95% Credible Interval (Cr. I) 1.06 – 1.66, see Supplementary Table 3). The probability of MRSA infection was found to be higher in adults (OR 1.06, 95% Cr. I 0.82-1.39), and lower in senior adults (OR 0.66, 95% Cr. I 0.45-0.96), when compared to children. Notably, the first half of the year showed the highest odds of MRSA infection among adult patients (refer to Supplementary Table 3 for more details)

### Spatial risk of MRSA by remoteness

Remoteness index was associated with an increased odds of MRSA infection across northern Australia. Compared to inner regional, the average odds ratio of 1.22 (95% Cr. I 0.45-3.20) in outer regional; and average odds ratio of 2.18 (95% Cr. I 0.72-6.38) in remote regions; and an average odds ratio of 1.86 (95% Cr. I 0.63-5.41) in very remote regions. The statistically significant time points for MRSA and remoteness index were February 2016, March 2016, July 2016, October 2017, December 2017, May 2018 and August 2021(refer to Supplementary Table 4 for more details).

### Spatial risk of MRSA by socio-economic disadvantage

We did not find a statically significant association between MRSA and any of the SEIFA indices assessed (Index of Economic Resources, Index of Education and Occupation, Index of Relative Social Advantage and Disadvantage, or Index of Relative Social Disadvantage).

### Using Bayesian spatial models to enhance insights gained from routine surveillance data for MRSA

To demonstrate the effectiveness of Bayesian spatial models to improving raw surveillance **(Figure 5)**, we used monthly MRSA data from June 2021. Our model incorporated the tested significant risk factors such as age, sex and remoteness **(Figure 6).**

**Figure 5.**
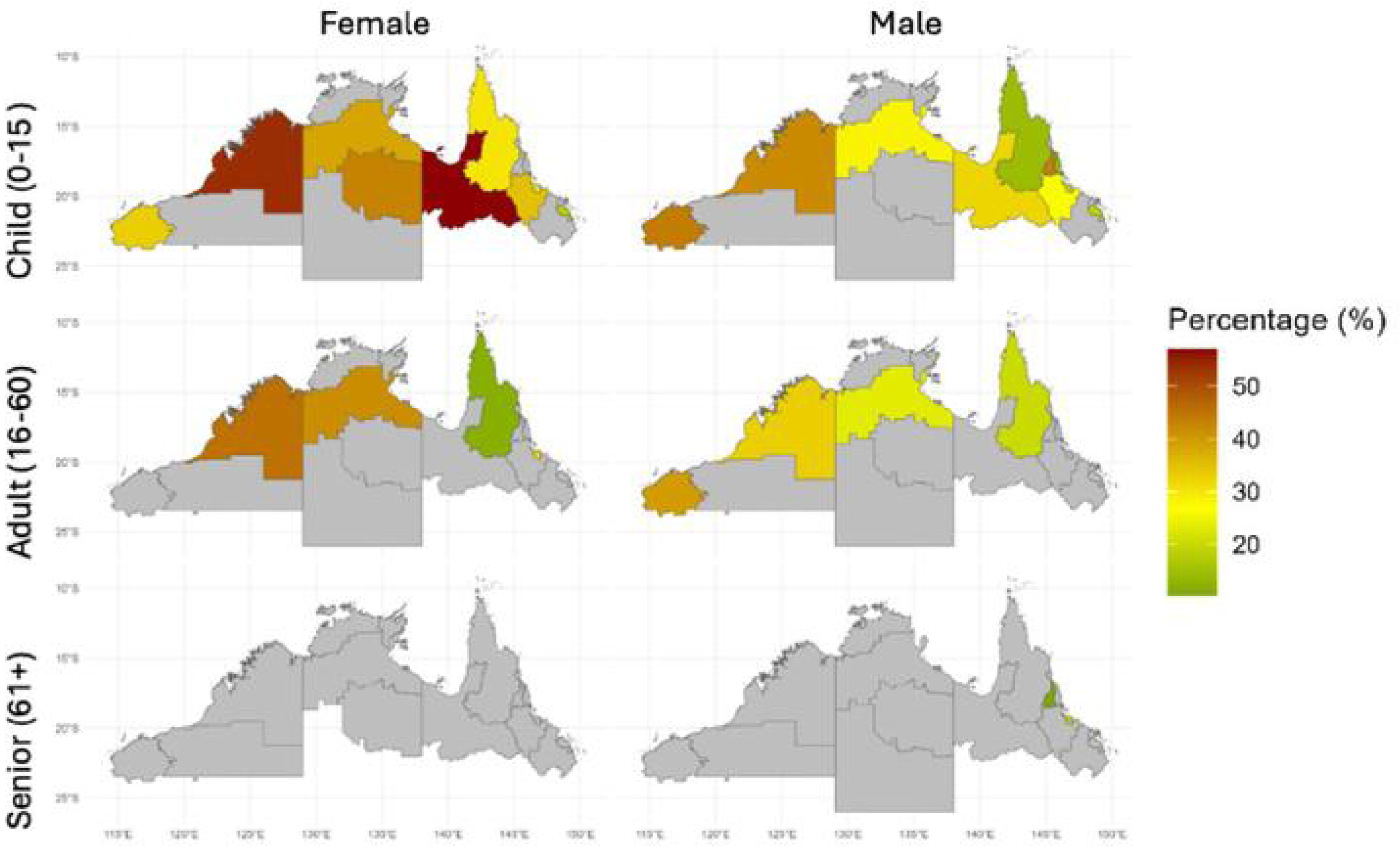
Map representing MRSA raw surveillance data by age and sex, northern Australia June 2021. Grey denotes regions where the total number of susceptibility tests was 15 or less for June 2021. No data was supplied for June 2021 from Alice Springs, Northern Territory (blank space for senior females).

**Figure 6.**
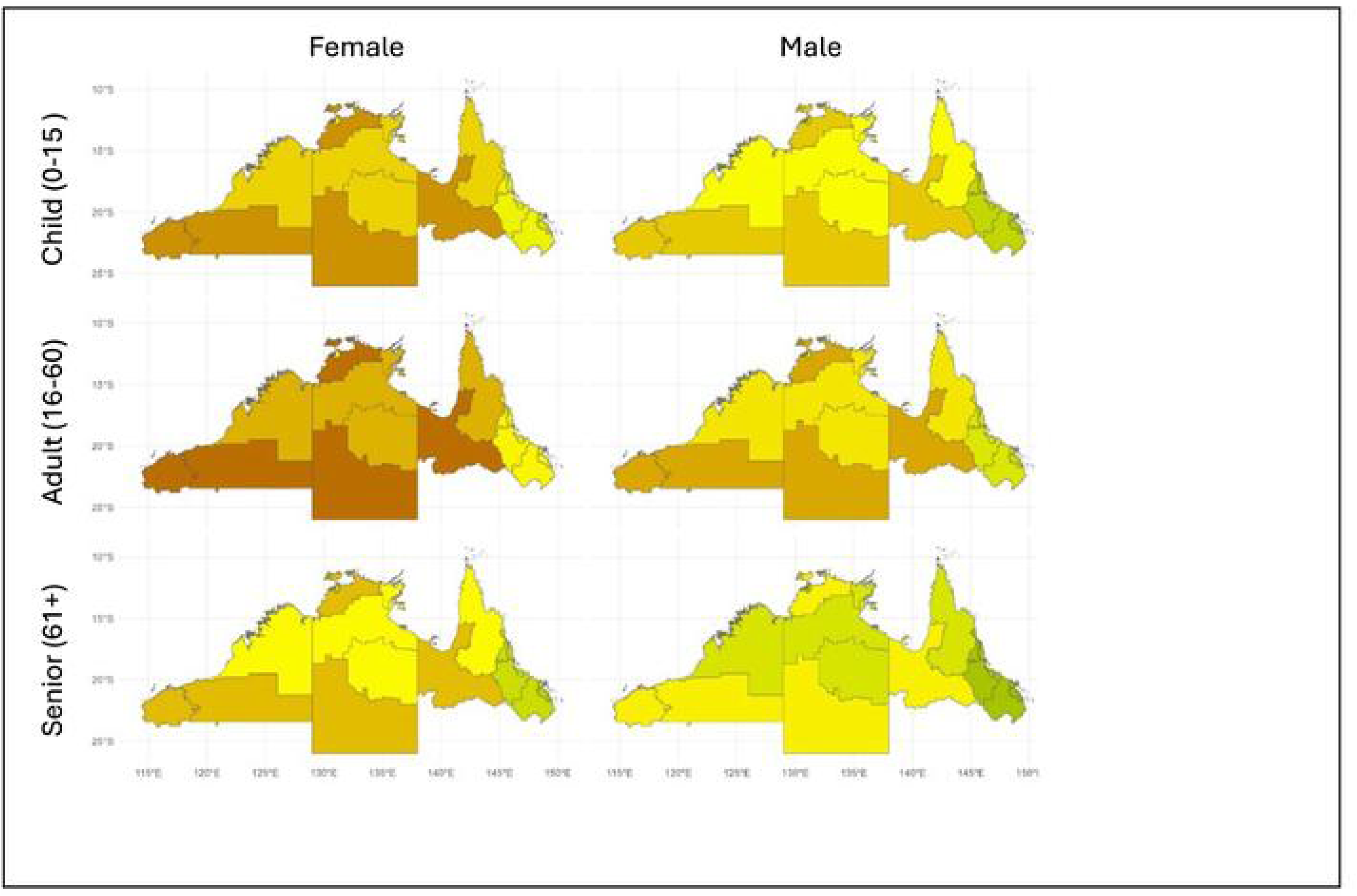
Map representing Bayesian spatial modelling to estimate posterior MRSA prevalence by age and sex, northern Australia June 2021.

In June 2021, when mapping raw surveillance data for each region by age and sex, it was found that 16 out of the total 22 geographical regions had missing data. This was due to either areas being unsampled or incomplete due to low counts (<15 records) (**Figure 5**). Therefore, less than 30% of the MRSA data could be utilised, with no data available for seniors (61 years and above) and limited data for adults (16-60 years) and children (0-15 years) in June 2021 as shown in Figure 5.

In contrast, using Bayesian spatial models permitted an analysis of the entire 22 regions for the month of June 2021(**Figure 6, Figure 7**). The regions with highest predicted prevalence across all age groups and sexes, were the remote regions of northern Australia which includes two Western Australia regions (east and west Pilbara), two Northern Territory regions (Daly-West Arnhem and Alice Springs) and one Queensland region (Outback North) **(Figure 6)**. The regions with lowest predicted MRSA infections were situated in urban settings and included regions in Northern Territory (Darwin region and Palmerston regions), and regions in east Australia (Mackay, Whitsunday, Townsville, Cairns).

**Figure 7.**
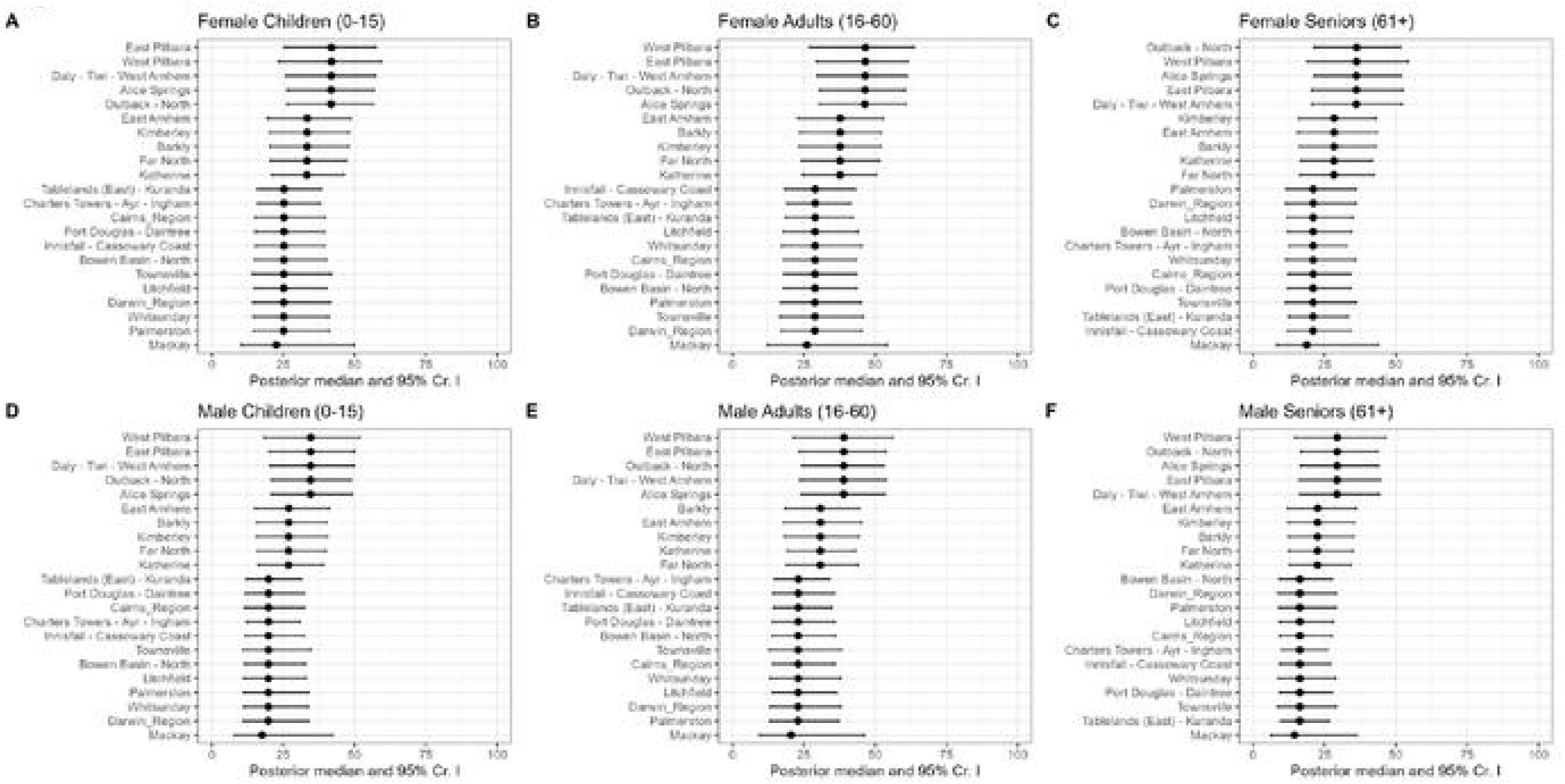
Posterior predicted MRSA prevalence estimate and 95% credible intervals by age and sex using Bayesian spatial models, northern Australia June 2021.

Posterior predicted estimates of MRSA and credible intervals are represented in Figure 7. Adult females (Figure 7B) and adult males (Figure 7E) were at highest risk of infection compared to children or seniors of each sex.

MRSA infection was highest in adult females (16-60 years) living in remote settings (West and East Pilbara regions:46.51% [95% Cr.I 29.34 – 61.72]), Daly-Tiwi-West Arnhem region: 46.43% [95% Cr. I 29.56 - 61.28], Alice Springs region: 46.51% [95% Cr. I 29.93-60.95], and Outback North region :46.45% [95% Cr.I 30.45-60.63]). MRSA infection was lowest in senior males (61+ years) living in urban settings (Mackay region 14.48%, and other regions including Townsville and Cairns).

## Discussion

We utilised Bayesian spatial modelling to generate monthly estimates of MRSA infections using routinely collected surveillance data in a sparsely populated, high-burden regions of Australia. From September 2020, we observed two distinct patterns of MRSA infection in northern Australia: potential clusters of high MRSA infections in the central regions, contrasted by low MRSA infection rates in the east coast regions. Our analysis revealed that MRSA was most prevalent amongst females aged 16-60 living in very remote areas, whereas senior males in urban areas of northern Australia had comparatively low MRSA infection rates.

These findings provide critical insights to refine our epidemiological understanding of MRSA rates within a region that has historically been beyond the reach of surveillance ^4^. We demonstrate the value of spatial modelling in pinpointing populations and geographic areas at increased risk of infection, as well as areas where infection rates remain low. This information will enable more targeted approaches to control AMR in high-risk regions and learn from regions that have a lower infection risk.

We identified large fluctuations in spatial autocorrelations in MRSA infections during the study period. From January 2017 to August 2020, spatial patterns of MRSA appeared random and highly variable. However, from September 2020, the patterns remained stable and did not change over time, suggesting a period of MRSA cluster formation. MRSA hot spots were observed in remote settings and central Australia regions, whilst cold spots were observed in urban settings and on largely east coast of Australia regions.

Potential for disease clustering likely has several underlying explanations and would merit further investigation. A primary driver for cluster formation starting from September 2020, could be attributed to the impact of coronavirus disease 2019 (COVID-19) pandemic. The pandemic not only resulted in significant loss of lives but also disrupted health systems and the economy to a considerable extent^24,25^. During the COVID-19 pandemic, Australia, like many other high income countries, experienced a decrease in the volume of antibiotic prescriptions^2^. Compared to 2019, this represented a substantial reduction, as much as 34% in the prescription of antimicrobials^2^. During the initial stage of the pandemic, there was a decrease in respiratory infections amongst children ^26^ and high uptake of enhanced hygiene and social distancing ^27^. However as the pandemic progressed, factors such as overuse of antibiotics in COVID-19 patients would have impacted the spread of AMR, with as many as 72% of COVID-19 patients receiving antibiotic treatment, either as empirical or to treat a bacterial coinfection, despite only 19% of these patients presenting with bacterial coinfections^28^. This would have been further compounded by the shortage of personal protective equipment, high staff turn-over and lack of healthcare staff throughout the pandemic ^29^. Additionally, changes in transmissibility or severity of MRSA, or the establishment of community-associated MRSA clones with specific virulence factors could have also been contributing factors^30^. This highlights the importance of improved and continuous surveillance efforts for MRSA, as fluctuations in incidence may indicate shifts in transmission dynamics or effectiveness of control measures.

We applied this spatial analysis to also investigate health inequality based on location. We found strong association between MRSA infection and remoteness, age and sex but not socio-economic factors, which supports previous findings^31^. The highest risk for MRSA infections was among adult females aged 16-60, living in very remote regions. Females had a 33% increased odds of MRSA infection compared to males (mean OR 1.33, 95% CI 1.06 – 1.66) across the study period. This may indicate a broader trend of higher resistance or vulnerability among females and could be due to various factors, including an observed effect in other settings of delayed antibiotic treatment in female patients^32^ or suggest a difference in health-seeking behaviours between the genders. The lowest risk was amongst senior males aged 60 years and over, living in inner regional areas, with a 44% reduced odds of MRSA infection (OR 0.66, 95% CI 0.45-0.96) compared to female seniors, adults and children as well as male adults and children. This groups lower risk of MRSA serves in contrast to the higher risk seen in other groups, indicating that factors such as age, sex, and location play a role in AMR. They underpin the need to understand how these factors reinforce AMR risk and vulnerability ^33^. This points to the need for targeted interventions, such as improving healthcare access in remote areas, ensuring timely antibiotic treatment and to consider age and sex-specific strategies to manage AMR infections.

The finding that MRSA infections in northern Australia were not associated with socio-economic status raises an important issue of using indices that measure individual level characteristics at a large geographical scale. We utilised the Australian Bureau of Statistics Statistical Area Level 3 (SA3) to determine socio-economic status. However, this approach may have obscured the impact on MRSA infection due to inherent variations in individual characteristics within such expansive administrative boundaries, particularly for residents of the Northern Territory and far north Western Australia, both of which cover vast geographical areas^34^. External analyses on the number of PBS prescriptions supplied for all antimicrobials per 1,000 individuals standardised by age at the SA3 level between 2017 to 2022 demonstrated a large range in the number of prescriptions across regions. Inner regional Mackay had a consistent 6-fold increase in the number of prescriptions compared to very remote East Arnhem over this period, indicating that in addition to potential socio-economic factors, antibiotic consumption rates may be an important factor to consider in future AMR modelling. However, due to the confidential nature of the PBS report at the time, we were unable to incorporate such information in our current analyses.

Inclusion of other data sources and understanding the needs of the population would also be critical in assessing spatial inequity^35^^36^. Bayesian modelling can overcome some of these challenges by integrating diverse data sources (with small sample sizes) and leveraging spatial-temporal dependence to refine and fill the information gaps for more meaningful and actionable epidemiological insights. Our estimates and maps of estimated MRSA prevalence show the fine detail of how age, sex and remoteness vary across space in a given month in these regions. We generate monthly MRSA rates by area which are more timely and provide spatial patterns for targeted AMR control efforts in a way that is not possible using raw unmodelled yearly surveillance data.

### Limitations

There are several limitations in this study. Firstly, the number of community healthcare clinics and hospitals participating in HOTspots aims to be comprehensive. However, there are several large private pathology services in Queensland not supplying data to HOTspots (e.g., Sullivan Nicolaides). Therefore, HOTspots surveillance data likely under-represent the private sector (community healthcare clinics) in far north Queensland. This is less of an issue in the NT and far north WA where Western Diagnostic Pathology services are the majority of the community healthcare clinics and contribute data to HOTspots. Due to the limited reporting of the exact location and number of private pathology services across northern Australia, it remains unclear what proportions of the entire northern Australian population are not included in the HOTspots surveillance system. However, as our data was aggregated to a region (i.e. Statistical Area Level 3), we are confident that each region would have a representative number of isolates from community healthcare clinics required for the analysis. Secondly, we used Bayesian spatial modelling to help improve the estimation of the statistical uncertainty of model parameters and smooth estimates, and to help generate better predictions at locations without data. However, there is the possibility for this approach to under-smooth or over-smooth estimates or predictions. Thus, we cannot discount the possibility that estimates of prevalence in particular region without any data might have been overestimated when lending weight from observed high prevalence values in a neighbouring location, or vice versa when there are excessive low or zero prevalence values in a neighbouring region. Thirdly, while our models converged in our spatial analyses of 22 geographical aerial regions, for replication of our methods in instances with sparse data or fewer regions, we recommend the implementation of zero inflated or sparse methods. Lastly, the spatial clustering of MRSA infections in our analysis suggests a potential for outbreaks in northern Australia but the Moran’ I method cannot quantify which regions within our dataset of northern Australia had similar MRSA infection rates. In future, a comprehensive assessment of widely employed cluster detection methods (i.e. Getis Ord G∗ i, Kulldorff spatial scan statistic) is recommended to identify the optimal cluster detection method for AMR surveillance data, as has been shown for dengue surveillance data^37^.

### Conclusion

The approach of spatial modelling and ‘borrowing’ information from neighbouring regions is especially needed to support AMR surveillance in regions that are resource poor, isolated or have diverse geographical populations. Integrating Bayesian spatial models into routine AMR surveillance systems, such as HOTspots, and developing further disease cluster detection methods for future analysis would be critical to identify the current and emerging high-risk areas and target these to reduce the continuous and worsening burden of AMR in northern Australia. This approach is flexible and scalable to other settings in Australia and beyond.

Finally, as temperatures continue to rise in northern Australia ^38^ such spatial modelling tools will be even more critical as the geographic distribution of infectious disease changes^39^ and the link between AMR and climate emerges ^40^. While system dynamics modelling will help understand the interactions and feedback loops ^41^of climate-related risks across One Health, spatial mapping using routine surveillance data will be critical to document these impacts across space and time.

## Data Availability

Models used for our study are available in the supplementary documents. Other data can be requested directly from authors, where appropriate.

## Authors’ contributions

TMW contributed to the conceptualisation, interpretation of results and writing and reviewing the manuscript. AD assisted with analytic interpretation and drafting of the final manuscript. AS and DC contributed to data cleaning, data aggregation, and review of the manuscript. AY contributed to modelling and generating all maps and tables in the manuscript. BS contributed with analytic interpretation and review of the manuscript. MC undertook data processing, development of the methodology, all modelling oversight, writing and reviewing the manuscript.

